# Risk of Cancer After Diagnosis of Cardiovascular Disease

**DOI:** 10.1101/2022.08.08.22278548

**Authors:** Caitlin F. Bell, Xiudong Lei, Richard Baylis, Hua Gao, Lingfeng Luo, Sharon H. Giordano, Mackenzie R. Wehner, Kevin T. Nead, Nicholas J. Leeper

## Abstract

**Background:** Cardiovascular disease (CVD) and cancer share several risk factors. While preclinical models show that various types of CVD can accelerate cancer progression, clinical studies have not determined the impact of atherosclerosis on cancer risk.

**Objectives:** To determine whether CVD, especially atherosclerotic CVD, is independently associated with incident cancer.

**Methods:** Using IBM MarketScan claims data from over 130 million individuals, we identified 27 million cancer-free subjects with a minimum of 36 months of follow-up data. Individuals were stratified by presence or absence of CVD, with 1:1 propensity matching to control for cardiovascular risk factors, and cumulative risk of cancer was calculated. Additional analyses were performed according to CVD type (atherosclerotic vs non-atherosclerotic) and cancer subtype.

**Results:** Among 4,487,412 matched individuals, those with CVD had a 1.26-fold higher relative risk of cancer than those without CVD (6.8% vs 5.4% 5-year cumulative incidence). Results were more pronounced for individuals with atherosclerotic CVD (aCVD), who had a 1.43-fold higher relative risk than those without CVD (7.7% 5-year cumulative cancer incidence). Findings persisted after multivariable adjustment for numerous traditional CV risk factors, including the modestly higher risk for cancer amongst individuals with atherosclerotic CVD. Cancer subtype analyses showed specific associations of aCVD with several malignancies, including lung, bladder, liver, brain, and other hematologic cancers.

**Conclusions:** Individuals with CVD have an increased risk of developing cancer compared to those without CVD. This association may be driven in part by the relationship of atherosclerosis with specific cancer subtypes, which persists after controlling for conventional risk factors.

## Main Text

Cardiovascular disease (CVD) and cancer continue to represent the two leading causes of death in the United States (US) (1, 2). These diseases share a multitude of risk factors, underscored by the reduction in mortality from both conditions when patients adhere to cardiovascular risk reduction guidelines (3-5). However, it is becoming increasingly clear that the two diseases may have a more complicated relationship, including shared pathophysiological mechanisms that extend beyond traditional risk factors.

Recent preclinical studies using murine models of heart failure, cardiac remodeling, or myocardial infarction have each demonstrated that solid tumors grow more quickly in the presence of cardiovascular abnormalities (6-8). In addition, several epidemiologic studies have shown that various forms of CVD may be associated with increased cancer progression and reduced survival (6-9). However, these retrospective cohort studies have been unable to determine which forms of CVD are associated with increased risk and, more importantly, if this relationship is simply driven by shared risk factors. Further, it remains unclear whether there is increased risk across all cancer types or if it is specific for certain cancers (5, 10, 11).

The current study aims to investigate the association of CVD, both atherosclerotic and non-atherosclerotic, with the development of cancer. An understanding of this relationship has the potential to better inform cancer risk stratification and screening (12, 13). In addition, improved characterization of the interaction between cancer and CVD could ultimately guide further mechanistic efforts to investigate pathophysiology mediating their interaction.

## Methods

### Data Sources

We performed a retrospective cohort study using data from the 2009-2019 IBM MarketScan® Research Databases (IBM, Ann Arbor, MI), which contains de-identified data for approximately 161 million patients including enrollment records and health insurance claims from inpatient services, outpatient visits, and outpatient prescription drugs. This study followed the Strengthening the Reporting of Observational Studies in Epidemiology (STROBE) reporting guidelines for cohort studies and was exempted by the University of Texas MD Anderson Cancer Center Institutional Review Board because of its use of deidentified data.

We additionally linked individuals who responded to the health risk assessment (HRA) survey in the 2009-2019 MarketScan Databases (IBM Corp, Armonk, NY) through unique enrollee identifiers. The HRA data contains biometric and behavioral information collected from risk assessment questionnaires administrated by the US corporations and health plans that contributed data to MarketScan. Specifically, it provided self-reported information on health indicators such as body mass index (BMI) and smoking status. It has been shown that the prevalence estimates of both variables are comparable to national estimates (14-16).

### Study Design

In this retrospective cohort study, we included patients 18 years of age or older enrolled since 2009 and with at least 36 months of continuous enrollment, and no cancer diagnosis codes in the first 24 months of enrollment. This yielded a cohort of over 27.5 million individuals. After a 24-month run-in period, these were classified into two groups depending on the presence or absence of CVD. Presence of CVD was defined as two or more separate *International Classification of Disease (ICD)*, Ninth (ICD-9) or Tenth (ICD-10) edition diagnosis or *Current Procedural Terminology* (*CPT*) codes indicating either ischemic or nonischemic CVD, in either the inpatient or outpatient setting (Supplemental Table 1). Codes were selected from previously validated and utilized algorithms demonstrating a >95% specificity of single code use for coronary artery disease, peripheral arterial disease, cerebrovascular disease, heart failure, valvular disease, congenital heart disease, (17-28), and >85% specificity of single code use for atrial fibrillation (29). Our study then requires two or more separate diagnostic codes to further increase specificity of our enrollment groups. Individuals without CVD and those with CVD were then matched based on a propensity score with an optimal caliper width that was 0.2 times the pooled standard deviation of the propensity score logit. The propensity score was derived from a logistic regression model where the independent variable was CVD (yes vs. no) and the covariates included age, sex, and presence of diabetes, hypertension, chronic kidney disease, and hyperlipidemia. The primary outcome of interest was incident cancer, defined as two or more separate cancer ICD-9/ICD-10 codes after the 24-month run-in period, either in the inpatient or outpatient setting (Supplemental Table 1). The index date was defined as 24 months after the date of first enrollment when the comparison groups were defined. Non-melanoma skin cancers were not included in either enrollment or outcome metrics.

As a secondary analysis, we utilized individuals with HRA-linked data to adjust our models for self-reported BMI and smoking status (n=1,282,261), in addition to age, sex, and presence of diabetes, hypertension, chronic kidney disease, and hyperlipidemia. As a separate sensitivity analysis, we excluded heart failure codes from the definition of CVD given data supporting an epidemiologic association between heart failure and incident cancer (10).

### Statistical Analysis

Groups were compared using a Chi-square test. Baseline characteristics included first enrollment year, age, sex, baseline presence of diabetes, hypertension, chronic kidney disease, and hyperlipidemia as well as geographic region and insurance type. The primary outcome was defined as the time from index date to the date of any cancer incidence (event) or last follow-up date (censor, end of enrollment or end of study December 31, 2019). Secondary outcomes included time from index date to the diagnosis of each of the top 20 most frequent organ-specific cancers (event) or last follow-up date (censor). For the primary outcomes, we implemented Kaplan-Meier estimates with log-rank test and multivariable Cox proportional hazards (PH) regression models to examine the association of groups with any cancer incidence, adjusting for all baseline characteristics and the propensity score, as derived for matching. We conducted a sensitivity analysis using a more conservative model with inverse probability treatment weighted (IPTW) based on propensity score (PS) with the weights being PS/(1-PS) for CVD groups and 1 for no CVD group.

For secondary outcomes, we evaluated the cumulative incidence of each of the top 20 most frequent cancers via a cause-specific approach. For example, when evaluating time to lung cancer incidence, patients who developed other organ-specific cancers were censored. We implemented cause-specific Cox PH regression model to evaluate the association of CVD groups with each incident cancer subtype outcome. We checked the proportional hazards assumption using Schoenfeld residuals.

We considered a two-sided p-value < 0.05 statistically significant unless otherwise indicated. When examining cancer subtypes, we declared statistical significance by setting a false discovery rate (FDR) q-value at 0.05 using the Benjamini-Hochberg approach to adjust for multiple comparisons. Individuals with missing data for each variable were treated as a separate unknown category with no imputation. All statistical analyses were performed using SAS Enterprise Guide version 9.4 (SAS Institute) and R 4.0.5 (R Foundation for Statistical Computing).

## Results

### Risk of All CVD

A total of 4,487,412 individuals with or without CVD underwent 1:1 matching for analysis (Figure 1). The mean age was 56 ± 16 years and 48% were female. Medical comorbidities included diabetes (25%), hypertension (65.6%), chronic kidney disease (2.8%), and hyperlipidemia (62.7%). The median follow-up time was 34 months ranging from 12 to 120 months. Table 1 summarizes cohort baseline characteristics. All available cardiovascular risk factors and co-morbidities including age, sex, and diagnosis of hypertension, diabetes, dyslipidemia, and renal failure were fully matched across the cohorts.

**Figure 1.**
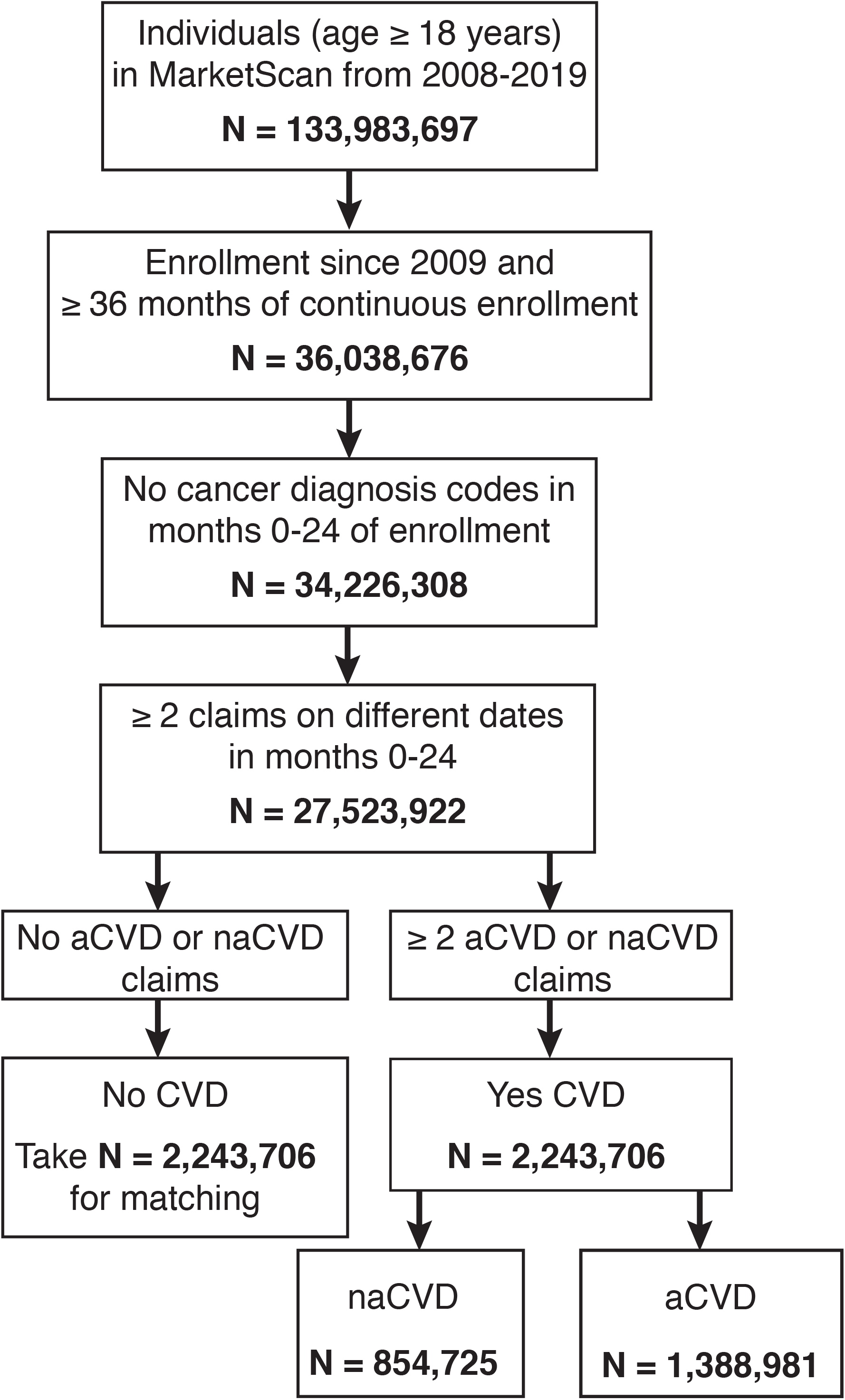
Cohort selection from MarketScan databases. Abbreviations: CVD, cardiovascular disease; naCVD, non-atherosclerotic cardiovascular disease; aCVD, atherosclerotic cardiovascular disease.

**Central Illustration.**
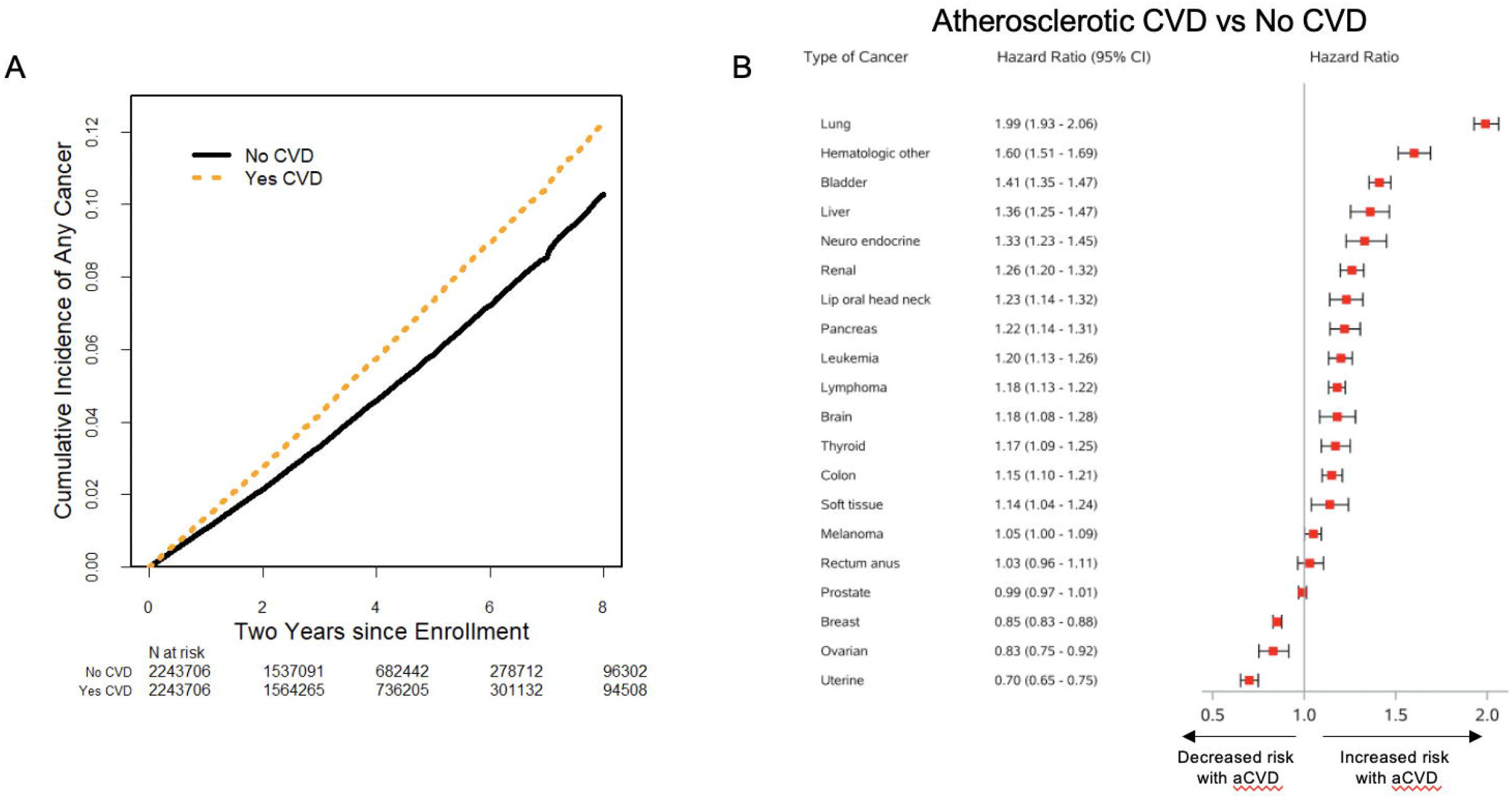
CVD, cardiovascular disease. (A) Kaplan-Meier estimates of time from index date to any cancer diagnosis among 1:1 matched cohorts (N=4,487,412) according to cardiovascular disease (CVD) group. Index date was set at 24 months after the date of first enrollment. (B) Forest plot of adjusted hazard ratios based on Cox proportional hazards model of time to organ-specific cancer incidence among 1:1 matched cohorts (N=4,487,412). All p values < 0.001 except p = 0.005 for soft tissue, p = 0.04 for melanoma, and and > 0.05 for both ovarian and uterine cancer. All multivariable Cox proportional hazards models were additionally adjusted for age, sex, baseline diabetes, chronic kidney disease, hyperlipidemia, region, and insurance.

**Table 1.** Baseline characteristics of 1:1 matched cohorts by cardiovascular (CVD) group

|  | No CVD<br>(N=2,243,706) | CVD<br>(N=2,243,706) | P Value |
| --- | --- | --- | --- |
|  | N (%) | N (%) |  |
| Age at first enrollment |  |  |  |
| Mean(std) | 54 (17) | 58 (15) |  |
| Median (IQR) | 56 (45, 65) | 58 (49,68) |  |
| 18-39 | 428348 (19.1) | 237459 (10.6) | <0.001 |
| 40-49 | 327274 (14.6) | 335055 (14.9) |  |
| 45-59 | 616422 (27.5) | 668658 (29.8) |  |
| 60-64 | 279451 (12.5) | 298706 (13.3) |  |
| 65-69 | 182814 (8.1) | 204977 (9.1) |  |
| 70-79 | 281483 (12.5) | 329910 (14.7) |  |
| ≥80 | 127914 (5.7) | 168941 (7.5) |  |
| Female | 1075928 (48) | 1075928 (48) | 1 |
| Diabetes | 549814 (24.5) | 570905 (25.4) | <0.001 |
| Hypertension | 1471549 (65.6) | 1471549 (65.6) | 1 |
| CKD | 63862 (2.8) | 60104 (2.7) | <0.001 |
| Hyperlipidemia | 1407731 (62.7) | 1407731 (62.7) | 1 |
| Region |  |  |  |
| Northeast | 462972 (20.6) | 568413 (25.3) | <0.001 |
| North central | 530317 (23.6) | 529578 (23.6) |  |
| South | 802216 (35.8) | 793443 (35.4) |  |
| West | 393225 (17.5) | 297275 (13.2) |  |
| Unknown | 54976 (2.5) | 54997 (2.5) |  |
| Insurance |  |  |  |
| PPO | 1244271 (55.5) | 1246496 (55.6) | <0.001 |
| HMO | 321499 (14.3) | 270061 (12) |  |
| Other | 552088 (24.6) | 589761 (26.3) |  |
| Unknown | 125848 (5.6) | 137388 (6.1) |  |

Abbreviations: CVD, cardiovascular disease; IQR, interquartile range; CKD, chronic kidney disease; PPO, preferred provider organization; HMO, health maintenance organization.
Note: All P-values comparing no CVD vs. CVD were significant with $p < 0.001$ except for sex, hypertension and hyperlipidemia.

In total, 3.67% of individuals without CVD compared to 4.75% of those with CVD developed incident cancers in the matched cohort. Incidence rates per 1000 person-years were 11.9 and 14.9 for the no CVD and CVD groups, respectively. An unadjusted estimate of 5-year cumulative incidence of cancer diagnosis in the matched cohort was 6.8% (95% CI, 6.7%-6.8%) for the CVD group compared to 5.4% (95% CI, 5.3%-5.4%) for the no CVD group (log-rank P<0.001) (Central Illustration). Multivariable and propensity score adjusted Cox PH modeling revealed that individuals with CVD were at significantly higher risk of developing incident cancer (HR, 1.14; 95% CI, 1.13-1.15) (Table 2), which remained consistent after sensitivity analysis using Cox PH models with IPTW (HR, 1.11; 95% CI, 1.10-1.13).

**Table 2.**
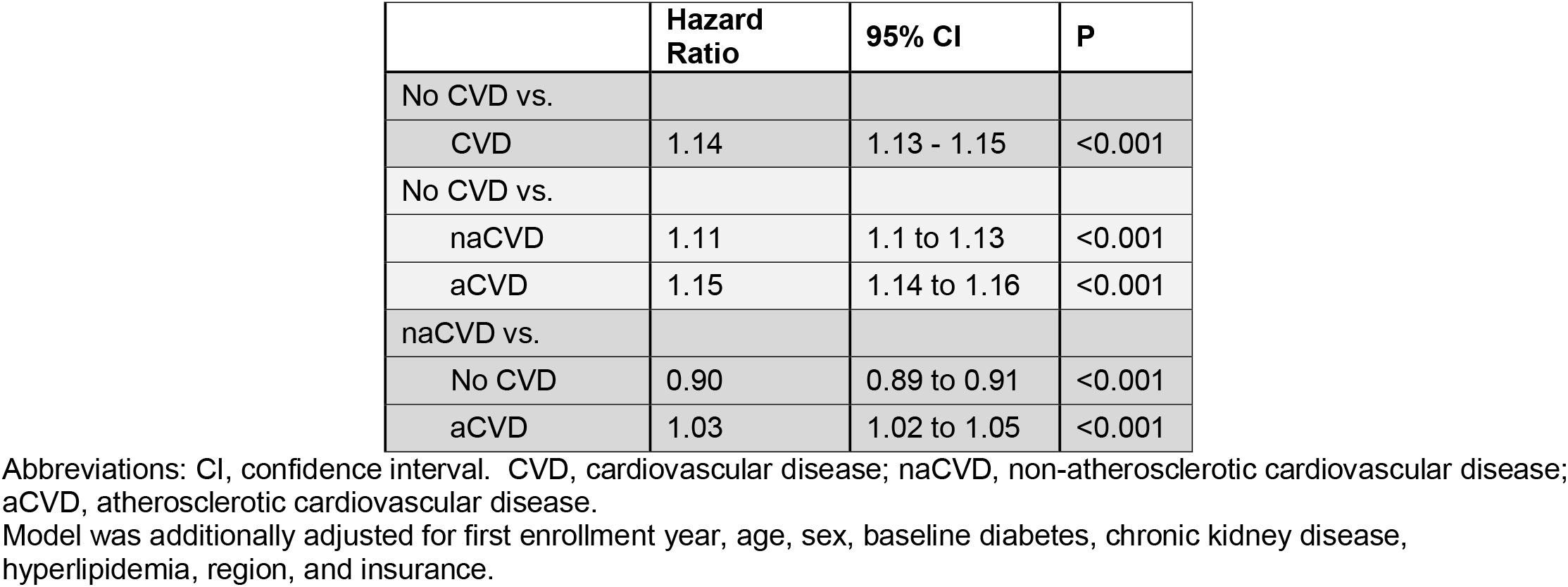
Cox proportional hazards model of time to any cancer incidence among 1:1 matched cohorts with propensity score adjustment (n=4,487,412)

|  | **Hazard Ratio** | **95% CI** | **P** |
| --- | --- | --- | --- |
| No CVD vs. |  |  |  |
| CVD | 1.14 | 1.13 - 1.15 | <0.001 |
| No CVD vs. |  |  |  |
| naCVD | 1.11 | 1.1 to 1.13 | <0.001 |
| aCVD | 1.15 | 1.14 to 1.16 | <0.001 |
| naCVD vs. |  |  |  |
| No CVD | 0.90 | 0.89 to 0.91 | <0.001 |
| aCVD | 1.03 | 1.02 to 1.05 | <0.001 |

Abbreviations: CI, confidence interval. CVD, cardiovascular disease; naCVD, non-atherosclerotic cardiovascular disease; aCVD, atherosclerotic cardiovascular disease.
Model was additionally adjusted for first enrollment year, age, sex, baseline diabetes, chronic kidney disease, hyperlipidemia, region, and insurance.

### Risk of Non-atherosclerotic and Atherosclerotic CVD

The CVD group was then split into aCVD and naCVD. The aCVD cohort was older with a higher proportion of men and medical comorbidities compared to the naCVD cohort (Supplemental Table 2).

5.46% of individuals with aCVD developed incident cancer compared to 3.60% of those with naCVD and 3.67% of those without CVD. Incidence rates per 1000 person-years were 17.5 and 11.4 for aCVD and naCVD groups, and 11.9 for the no CVD group. The 5-year cumulative incidence was 5.4% (95% CI, 5.3%-5.4%) for aCVD and 5.2% (95% CI, 5.2%-5.3%) for the naCVD group (Figure 2), compared to 1.16% (95% CI, 1.12%-1.19%) for the no CVD group (log-rank p <0.001).

**Figure 2.**
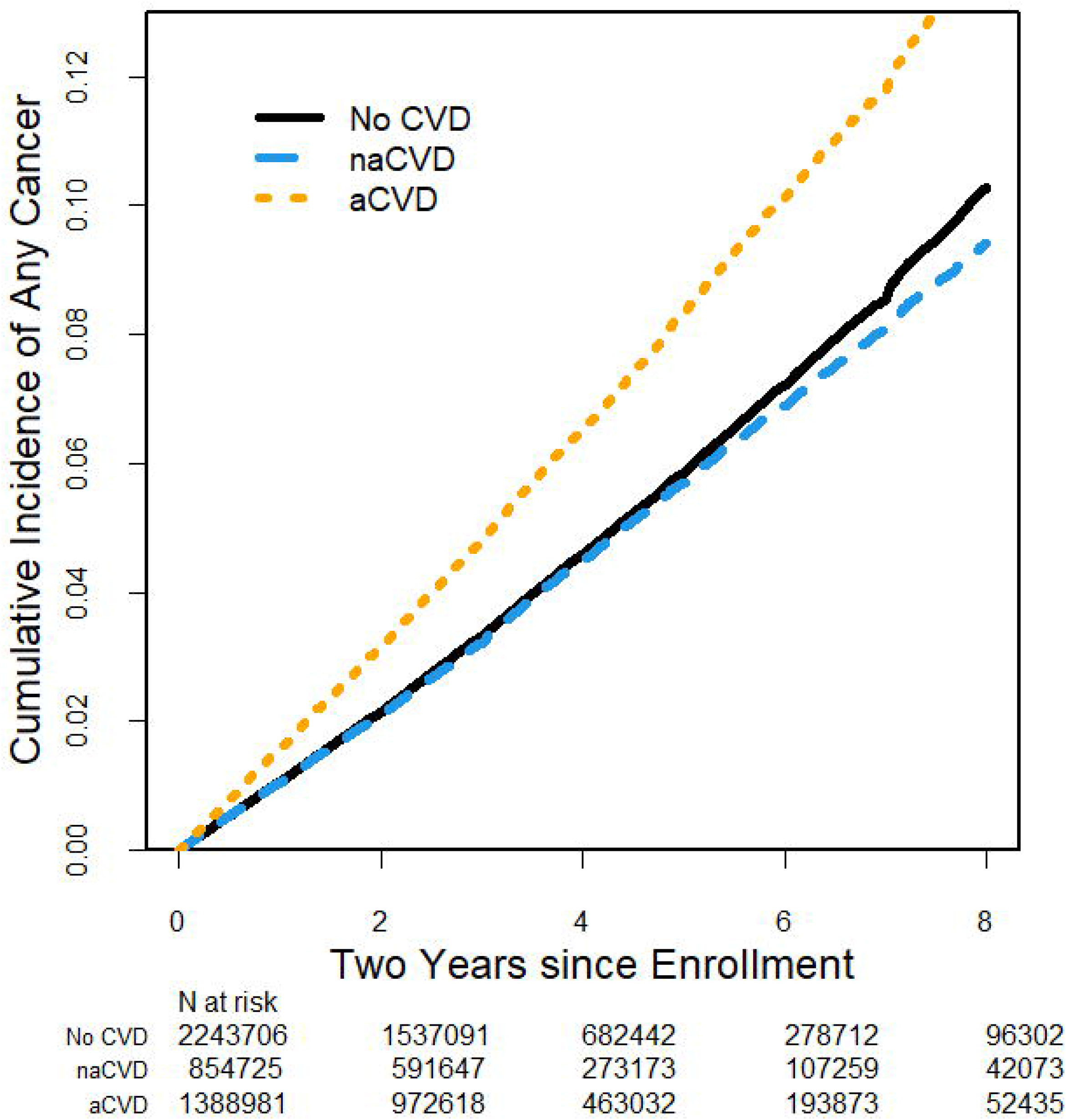
Cumulative incidence of cancer according to cardiovascular disease group. Abbreviations: CVD, cardiovascular disease; naCVD, non-atherosclerotic cardiovascular disease; aCVD, atherosclerotic cardiovascular disease. Kaplan-Meier estimates of time from index date to any cancer diagnosis among 1:1 matched cohorts (N=4,487,412) according to cardiovascular disease (CVD) group. Index date was set at 24 months after the date of first enrollment.

To control for imbalances in age, sex and other comorbidities between those with atherosclerotic versus non-atherosclerotic forms of CVD, multivariable adjusted Cox proportional hazards models were generated. These studies revealed that those with aCVD (HR, 1.15; 95% CI, 1.14-1.16) and naCVD (HR, 1.11; 95% CI, 1.10-1.13) were both at significantly increased risk of incident cancer compared to individuals without CVD (Table 2). In addition, the risk of incident cancer remained modestly but statistically significantly higher in the aCVD group when directly compared to the naCVD group (HR, 1.03; 95% CI, 1.02-1.05) in these analyses. Cox PH models with IPTW revealed a consistent result when comparing individuals with aCVD (HR, 1.12; 95% CI, 1.10-1.13) and naCVD (HR, 1.10; 95% CI, 1.08-1.12) to those with no CVD.

To determine whether our results were being driven by an association of heart failure and cancer, we conducted a sensitivity analysis excluding all individuals with heart failure diagnosis codes. Results were consistent with our primary analysis demonstrating an increased risk of cancer among those with CVD (HR: 1.13; 95% CI, 1.12-1.14), and specifically, aCVD (HR: 1.14; 95% CI, 1.13-1.15) and naCVD (HR: 1.11; 95% CI, 1.09-1.12) compared to those without CVD after multivariable and propensity score adjustment which controlled for available cardiovascular comorbidities.

Analysis among those with HRA data, providing additional adjustment for self-reported smoking status and BMI (14-16), included 1,282,261 individuals. The cohort was propensity score matched for all available CV risk factors, including smoking status and BMI. Cohort characteristics are summarized in Supplemental Table 3.

The HRA-linked data similarly demonstrated that unadjusted estimates of 5-year cumulative cancer incidence were significantly higher in the CVD group 4.0% (95% CI, 3.8%-4.1%) than those without CVD 2.1% (95% CI, 2.1%-2.2%, log-rank p<0.001). Furthermore, the accentuation of risk according to the presence or absence of atherosclerosis persisted, including rates of 4.6% (95% CI, 4.3%-4.9%) in the aCVD group and 3.3% (95% CI, 3.1%-3.6%) in the naCVD group. These results were consistent in a multivariable adjusted Cox PH model incorporating smoking status and BMI (Supplemental Table 4). Compared to individuals without CVD, those with CVD were at significantly increased risk of developing cancer (HR, 1.16; 95% CI, 1.11-1.21). When those without CVD were compared to CVD subgroups, both aCVD (HR, 1.18; 95% CI, 1.12-1.25) and naCVD (HR, 1.13; 95% CI 1.06-1.2) had increased risk of incident cancer. The difference between naCVD and aCVD in this dataset was directionally consistent with our primary analysis albeit with lower power (HR, 1.05; 95% CI 0.97-1.14).

### Cancer Subtype Analysis

We used multivariable adjusted Cox PH models to examine the association of aCVD and naCVD with the twenty most frequently diagnosed cancers in our dataset (Central Illustration and Figure 3). Both aCVD and naCVD had significantly increased risk of multiple cancer subtypes compared to those without CVD, though the risk among multiple subtypes was more prominent in the aCVD group. aCVD was also noted to have a significantly lower risk for breast, uterine, and ovarian cancers. When directly compared, aCVD had a significantly higher risk than naCVD for cancers of the lung, bladder, colon, head and neck, liver, and brain as well as leukemia and other hematologic malignancies (Supplemental Figure 1).

**Figure 3.**
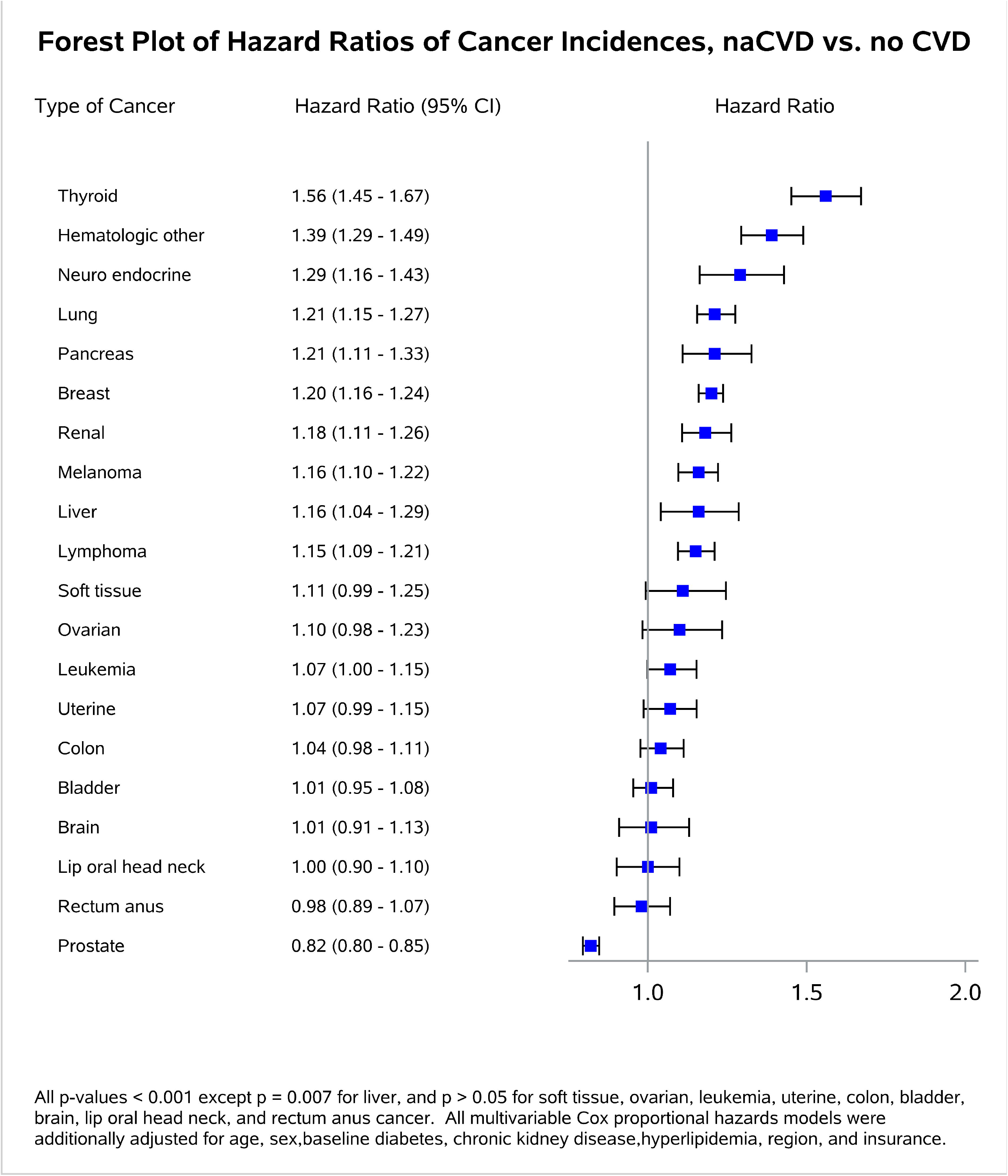
Abbreviations: CVD, cardiovascular disease; naCVD, non-atherosclerotic cardiovascular disease. Forest plots of adjusted hazard ratios based on Cox proportional hazards model of time to organ-specific cancer incidence among 1:1 matched cohorts (N=4,487,412).

Since several of these malignancies have known associations with tobacco use, we utilized our HRA-linked dataset to further control for smoking status and BMI in a multivariable analysis (Supplemental Figure 2). This analysis continued to observe an increased association for lung cancer (HR; 1.91, 95% CI; 1.55-2.36), bladder cancer (HR; 1.65, 95% CI; 1.30-2.09), as well as cancers not strongly associated with smoking such as other hematologic malignancies (HR; 1.68, 95% CI; 1.28-2.21), and lymphoma (HR; 1.48, 95% CI; 1.21-1.80), in the aCVD group compared to the no CVD group.

## Discussion

Our retrospective cohort study demonstrates that individuals with CVD have a significantly increased risk of incident cancer compared to those without CVD. We found that this risk was most pronounced amongst individuals with atherosclerotic CVD, even after adjusting for all available cardiovascular comorbidities. With data from over 4.4 million individuals, this is the largest study to explore this relationship. Importantly, our findings were consistent in a secondary analysis which controlled for self-reported smoking status and BMI, as well as when heart failure diagnoses were excluded, suggesting that the presence of heart failure or traditional CV risk factors may not fully account for the observed associations. Finally, cancer subtype analyses demonstrated specific associations between aCVD and a range of individual cancers, highlighting areas for future mechanistic research.

Our findings build upon prior studies. A study of 32,095 individuals from the Sakikibara Heart Institute in Japan comparing cancer incidence between those with atherosclerosis and those with nonischemic forms of CVD found the presence of atherosclerosis to be an independent risk factor for cancer diagnosis (11). Generalizability of these results, however, was limited by the absence of a healthy control group, limited study size to explore cancer subtypes, and a homogenous sample with a single ancestry of participants from a tertiary-care center. Our results build upon these findings by incorporating a healthy control group, increased power to determine cancer subtypes, and use of a more diverse patient population. Another analysis that utilized data from 20,305 Framingham Heart Study and PREVEND study participants found that traditional CV risk factors are associated with increased cancer incidence (5). This study, like the ARIC (Atherosclerosis Risk in Communities) and MESA (Multiethnic Study of Atherosclerosis) studies, showed that measures taken to improve cardiovascular health also decrease cancer risk (3, 4). Analysis of this cohort, however, did not ultimately demonstrate an increased risk of cancer diagnosis associated with CV events or CV prevalence, and was unable to determine whether the link was driven by underlying shared risk factors or the existence CVD itself (5, 30).

Two previous retrospective cohort studies found a connection between heart failure and cancer incidence (6, 10). These studies, however, did not distinguish between ischemic and nonischemic etiologies of heart failure and were unable to control for smoking. Our sensitivity analysis excluding heart failure diagnoses entirely from the dataset did not alter findings of our primary analysis; both the atherosclerotic and non-atherosclerotic groups continued to have increased cancer incidence compared to those without CVD. Therefore, our results do not appear to be driven by a specific association between heart failure and cancer.

The individual cancer analyses demonstrate that many cancer subtypes occur at higher rates in patients with CVD. The risk differed, however, according to the presence or absence of atherosclerosis. For example, in both the primary analysis and the secondary HRA-based analysis (which adjusted for BMI and smoking, in addition to age, sex, and each of the co-morbidities listed in Table 1), individuals with aCVD had an increased risk of lung cancer, bladder cancer, and certain hematologic malignancies relative to those with naCVD and those without CVD. Interestingly, traditionally hormone-driven cancers including breast and ovarian cancer demonstrated an inverse association, with aCVD conferring a protective effect. While mechanisms underlying this interesting association are unclear, it is interesting to note that estrogen impacts vasodilation and vascular remodeling and has been shown to have a net antiatherosclerotic effect (31). Additionally, aromatase inhibitors, which cause systemic estradiol depletion, have been associated with a modest increase in cardiovascular events in placebo-controlled RCTs (32, 33), There is also increasing recognition of several known shared pathways in sterol/oxysterol and hormone metabolism that influence plaque progression and hormone-sensitive malignancies (34).

It is enticing to hypothesize that these links between CVD and specific cancer subtypes are mediated through shared biologic processes (e.g., inflammation, metabolic adaptations, etc). Indeed, recent mouse studies have sought to explore the pathologic crosstalk between cancer and CVD. Two studies of heart failure models have found increased rates of cancer formation mediated by secreted factors (6, 7). Another using a murine model of acute myocardial infarction showed increased cancer progression mediated through innate immune system reprogramming (8). One study employing a mouse model of obesity/metabolic syndrome found that tumor growth was enhanced via alterations in T cells within the tumor microenvironment (35). While no preclinical studies have yet modeled the impact of atherosclerosis on tumorigenesis, these prior publications suggest that perturbations of cardiovascular homeostasis may promote tumorigenesis.

Human studies also suggest pathophysiologic overlap between cancer and CVD independent of shared traditional risk factors. For example, genome-wide association studies (GWAS) have shown that the most important commonly inherited genetic variant associated with atherosclerosis resides in a well-known cancer locus at chromosome 9p21, rather than in a gene which regulates traditional CV risk factors (36-40). Inflammation and immune cell activation have also been linked to both conditions. The CANTOS (Canakinumab Anti-inflammatory Thrombosis Outcomes) trial, designed to test the hypothesis that an anti-interleukin-1ß antibody could reduce adverse cardiovascular outcomes, surprisingly found a concomitant reduction in lung cancer and lung cancer mortality (41-44). Similarly, macrophage checkpoint inhibitors designed to reactivate anti-cancer immune surveillance machinery in subjects with lymphoma were recently found to simultaneously reduce vascular inflammation in those same individuals (45). A final area of overlap relates to the phenomenon of clonal hematopoiesis of indeterminate potential, (CHIP) which refers to the clonal expansion of mutated myeloid cells that can precede hematological malignancies. This process is also associated with risk for coronary disease, causing investigators to revisit the clonal hypothesis of atherosclerosis and its overlap with the clonally-expanding cancer stem cell. While the precise molecular mechanism linking CHIP mutations to heart disease remains undefined, inflammasome activation is thought to be central to this process (46-49). Interestingly, patients in the CANTOS trial with CHIP mutations benefited more from canakinumab than those without a CHIP mutation (49).

Our study has several limitations. First, our study design was observational in nature meaning causality cannot be demonstrated. Second, there is potential misclassification of diagnoses within the ICD coding system, though codes were chosen based on rigorous review of validated claims-based algorithms (17-29). We additionally required two separate diagnosis codes at two separate healthcare encounters to increase our confidence in how we defined exposure groups and cancer incidence. Third, our results may be subject to residual confounding from unknown covariables. While our secondary analysis using HRA data was able to control for BMI and smoking as covariables (14-16), this dataset was not able to account for cumulative pack-years. It also had fewer patients, forcing us to restrict our analysis to individuals between 18 to 65 years of age upon enrollment. Additionally, we were unable to control for certain other potential risk factors, such as physical activity, environmental exposures, alcohol consumption, and other sociodemographic variables. Finally, as our dataset does not include information on race or ethnicity it is impossible to discern whether these factors influence the relationship between CVD and cancer.

## Conclusions

This study found that the presence of CVD is associated with an increased incidence of cancer, particularly amongst individuals with atherosclerotic cardiovascular disease. Further work to understand the interaction between cardiovascular disease and cancer could have significant implications for the development of novel prevention, screening, and multidisciplinary therapeutic approaches for humanity’s two leading killers.

### Competency in Patient Care and Procedural Skills

Patients with CVD have disproportionately higher rates of incident cancer compared to those without CVD irrespective of shared traditional risk factors. The risk for certain types of malignancies varies depending on whether the patient has aCVD or naCVD.

### Translational Outlook

Further basic biological work is needed to understand the interactions of different CVD types with cancer, in addition to prospective studies considering whether patients with different types of CVD could benefit from adjusted cancer prevention or screening protocols.

## Data Availability

Data are available through the vendor on investigator application

## Abbreviation List

CVD: cardiovascular disease
aCVD: atherosclerotic cardiovascular disease
naCVD: nonatherosclerotic cardiovascular disease
HRA: health risk assessment
PS: propensity score

