## Supplemental Material for "Risk of Cancer After Diagnosis of Cardiovascular Disease"

Figure 1 – Forest plots of adjusted hazard ratios based on Cox proportional hazards model of time to organ-specific cancer incidence among 1:1 matched cohorts

Figure 2 – Forest plots of adjusted hazard ratios based on Cox proportional hazards model of time to organ-specific cancer incidence among HRA cohort

Table 1 – Diagnosis and procedure codes to define cardiovascular diseases, other comorbidities and cancer

Table 2 – Baseline characteristics of 1:1 matched cohorts by cardiovascular disease (CVD) group

Table 3 – Baseline characteristics of HRA-linked cohorts by cardiovascular disease (CVD) group

Table 4 - Cox proportional hazards model of time to cancer for HRA-linked data

Figure 1 – **Forest plots of adjusted hazard ratios based on Cox proportional hazards model of time to organ-specific cancer incidence among 1:1 matched cohorts** (N=4,487,412)

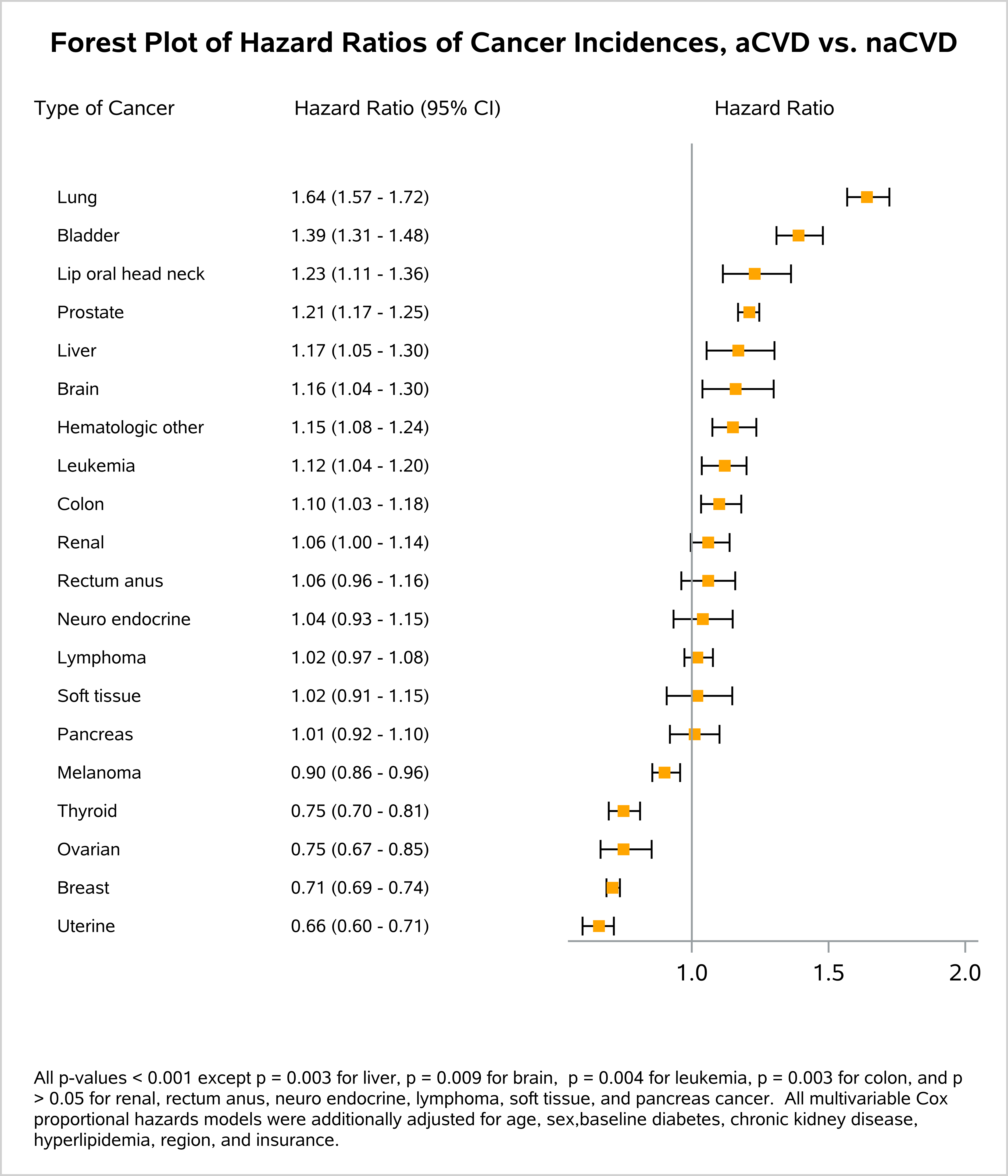

Abbreviations: CI, confidence interval. naCVD, non-atherosclerotic cardiovascular disease; aCVD, atherosclerotic cardiovascular disease.

Models were additionally adjusted for first enrollment year, age, sex, baseline diabetes, chronic kidney disease, hyperlipidemia, region, and insurance.

Figure 2 – **Forest plots of adjusted hazard ratios based on Cox proportional hazards model of time to organ-specific cancer incidence among HRA cohort** (N=1,282,261)

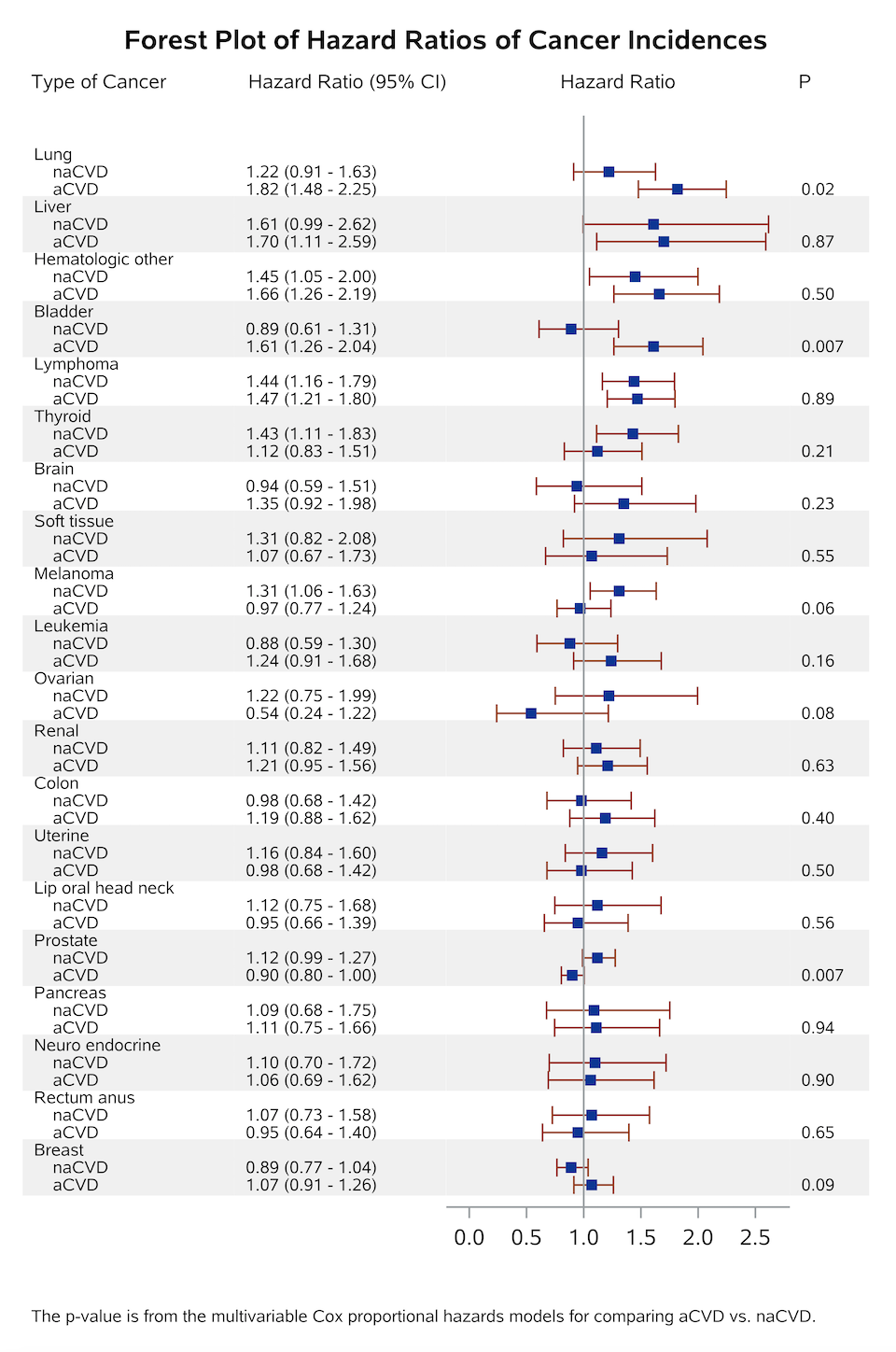

Abbreviations: CI, confidence interval. CVD, cardiovascular disease; naCVD, non-atherosclerotic cardiovascular disease; aCVD, atherosclerotic cardiovascular disease.

Models were additionally adjusted for first enrollment year, age, sex, baseline diabetes, chronic kidney disease, hyperlipidemia, region, insurance, Body Mass Index, and smoking status.

**Table 1. Diagnosis and procedure codes to define cardiovascular diseases, other comorbidities and cancer.**

| **Conditions** | **Diagnosis / Procedure Codes** | |
| --- | --- | --- |
| Atherosclerosis cardiovascular disease (aCVD) | Diagnosed with atherosclerotic cardiovascular disease (aCVD), defined as:   - Two of any of the following codes, diagnostic or procedural, on two separate inpatient or outpatient visits (e.g. a code listed under “coronary artery disease” and a code listed under “peripheral artery disease”, or two codes listed under “coronary artery disease”). - 413.x, I20.x (angina pectoris, ICD-9/-10 diagnosis) plus any of the following diagnostic or procedural codes, on two separate inpatient or outpatient visits (e.g. 413.x plus a code listed under “coronary artery disease”). - 425.xx, I43 (cardiomyopathy, ICD-9/-10 diagnosis) or 428.xx, I50.xxx (heart failure, ICD-9/-10 diagnosis) plus any of the following diagnostic or procedural codes, on two separate inpatient or outpatient visits (e.g. 425.xx plus a code listed under “coronary artery disease”). | |
|  | **Coronary artery disease** | Diagnosis  (ICD-9) 410.xx (acute myocardial infarction); 411.xx (other acute and subacute forms of ischemic heart disease); 412 (old myocardial infarction); 414.0x, 414.2 - 414.9 (coronary atherosclerosis); 429.7x (sequelae of myocardial infarction)  (ICD-10) I21.x (acute myocardial infarction; excluding I21.Ax); I22.x (STEMI/NSTEMI); I23.x (complications following STEMI/NSTEMI); I24.x (other acute ischemic heart disease); I25.x (chronic ischemic heart disease; excluding I25.3, I25.4x); Z95.5 (presence of coronary angioplasty implant and graft); Z98.61 (coronary angioplasty status).  T82.211, T82.211A, T82.211D, T82.211S, T82.212, T82.212A, T82.212D, T82.212S, T82.213, T82.213A, T82.213D, T82.213S, T82.218, T82.218A, T82.218D, T82.218S, Z951  Procedure  (ICD-9 CM) 0.66 (PCTA); 36.0x (removal of coronary artery obstruction); 36.1x (bypass anastomosis for heart revascularization); 36.2 (heart revascularization by arterial implants); 36.3x (other heart revascularization); 17.55 (transluminal coronary atherectomy).  (ICD-10 PCS Diagnosis) 0210xxx (coronary bypass, one artery); 0211xxx (coronary bypass, two arteries); 0212xxx (coronary bypass, three arteries); 0213xxx (coronary bypass, four arteries); 0270xxx (coronary dilation, one artery); 0271xxx (coronary dilation, two arteries); 0272xxx (coronary dilation, three arteries); 0273xxx (coronary dilation, four arteries)  (CABG) B2020ZZ, B2021ZZ, B202YZZ, B2030ZZ, B2031ZZ, B203YZZ, B212010, B2120ZZ, B212110, B2121ZZ, B212Y10, B212YZZ, B213010, B2130ZZ, B213110, B2131ZZ, B213Y10, B213YZZ, B22300Z, B2230ZZ, B22310Z, B2231ZZ, B223Y0Z, B223YZZ, B223Z2Z, B223ZZZ, B233Y0Z, B233YZZ, B233ZZZ  (CPT) (PCI) 92920, 92921, 92924, 92925, 92928, 92929, 92933, 92934, 92937, 92938, 92941, 92943, 92944, C9600, C9601, C9602, C9603, C9604, C9605, C9606, C9607, C9608  (CABG) 33510, 33511, 33512, 33513, 33514, 33516, 33517, 33518, 33519, 33520, 33521, 33522, 33523, 33525, 33528, 33530, 33533, 33534, 33535, 33536, 35600, 4110F, 75762, 75764, 75766, 75767, 93551, C9604, C9605, G8158, G8159, G8160, G8161, G8162, G8163, G8164, G8165, G8166, G8167, G8170, G8171, G8172, G8497, G8544, G8573, G8574 |
|  | **Peripheral artery disease, aortic atherosclerosis** | Diagnosis  (ICD-9) 433.xx (occlusion and stenosis of precerebral arteries); 440.xx (atherosclerosis); 443.9 (peripheral vascular disease, unspecified)  (ICD-10) I65.xx (occlusion and stenosis of precerebral arteries, not resulting in cerebral infarction); I70.xxx (atherosclerosis); I73.9 (peripheral vascular disease, unspecified); I75.xxx (atheroembolism)  Procedure  Peripheral Revascularization (carotid and cerebral included):  (ICD-9 CM) 00.55 (insertion of drug eluting non-coronary stent); 00.63 (perc insertion of carotid artery stent); 00.64 (perc insertion of extracranial artery stent); 00.65 (perc insertion of intracranial artery stent); 17.53 (perc atherectomy extracranial vessel); 17.54 (perc atherectomy intracranial vessel); 17.56 (atherectomy of noncoronary vessel); 38.1x (endarterectomy); 39.25, 39.26, 39.29 (peripheral bypass grafting); 39.50 (Angioplasty of other non-coronary vessel(s)); 39.90 (Insertion of non-drug-eluting peripheral (non-coronary) vessel stent(s)).  (ICD-10 PCS) 047Cxxx; 047Dxxx; 047Exxx; 047Fxxx; 047Hxxx; 047Jxxx; 047Kxxx; 047Lxxx; 047Mxxx; 047Nxxx; 047Pxxx; 047Qxxx; 047Rxxx; 047Sxxx; 047Txxx; 047Uxxx; 047Vxxx; 047Wxxx; 047Yxxx (lower extremities, dilation). 04C0xxx; 04CCxxx; 04CDxxx; 04CExxx; 04CFxxx; 04CHxxx; 04CJxxx; 04CKxxx; 04CLxxx; 04CMxxx; 04CNxxx; 04CPxxx; 04CQxxx; 04CRxxx; 04CSxxx; 04CTxxx; 04CUxxx; 04CVxxx; 04CY (extirpation of lower extremity artery). 04100Jx; 041C0Jx; 041D0Jx; 041H0Jx; 041H0Jx; 041H0Kx; 041H4Jx; 041J0Jx; 041J4Jx; 041K0Jx; 041K0Zx; 041L09x; 041L0Kx; 041L0Zx; 041Mxxx; 041Nxxx; 041Sxxx; 041Txxx; 041Uxxx (bypass of lower extremity artery).  (CPT) Peripheral Revascularization (carotid and cerebral included) 37205, 37206, 37207, 37208, 37236, 37237, 37184, 37185, 37186, 35302, 35303, 35304, 35305, 35306, 35331, 35351, 35355, 35361, 35363, 35371, 35372, 35381, 35452, 35454, 35456, 35459, 35470, 35472, 35473, 35474, 35483, 35492, 35493, 35495, 35521, 35533, 35537, 35538, 35539, 35540, 35556, 35558, 35563, 35565, 35566, 35571, 35583, 35585, 35587, 35621, 35623, 35637, 35638, 35646, 35647, 35654, 35656, 35661, 35663, 35665, 35666, 35671, 35700, 35876, 35879, 35881, 35883, 35884, 37184, 37185, 37186, 37205, 37206, 37207, 37208, 0236T, 0237T, 0238T, 37225, 37224, 37227, 37226, 37222, 37223, 37220, 37221, 37229, 37228, 37231, 37230, 37233, 37232, 37235, 37234, 35548 |
|  | **Cerebral vascular disease** | Diagnosis  (ICD-9) 433.xx (occlusion and stenosis of precerebral arteries); 434.x (occlusion and stenosis of cerebral arteries); 435.x (TIA); 437.x (cerebral atherosclerosis)  (ICD-10) G45.x (TIA); I63.xx (cerebral infarction); I65.xx (occlusion and stenosis of precerebral arteries, not resulting in cerebral infarction); I66.xx (occlusion and stenosis of cerebral arteries, not resulting in cerebral infarction); I67.2 (cerebral atherosclerosis) |
| Non-atherosclerosis CVD (naCVD) | Diagnosed with non-atherosclerotic CVD (naCVD), defined as:   - Two of any of the following codes, diagnostic or procedural, on two separate inpatient or outpatient visits. - If criteria for both aCVD and naCVD are met, then the patient will be placed in the aCVD group. | |
|  | **Myocardial diseases** | Diagnosis  (ICD-9) 398.xx (other rheumatic heart disease); 402.xx (hypertensive heart disease); 404.xx (hypertensive heart and chronic kidney disease); 415.0 (acute cor pulmonale); 416.8, 416.9 (chronic pulmonary heart disease); 422.xx (acute myocarditis); 425.xx (cardiomyopathy); 428.xx (heart failure); 429.0, 429.1, 429.3, 429.8x (other ill-defined heart diseases).  (ICD-10) I11.x (hypertensive heart disease); I13.x (hypertensive heart and chronic kidney disease); I25.3 (aneurysm of heart); I26.0x (pulmonary embolism with acute cor pulmonale); I27.xx (other pulmonary heart disease); I40.x (acute myocarditis); I42.x (cardiomyopathy); I43 (cardiomyopathy in diseases classified elsewhere); I50.xxx (heart failure); I51.xx (complications and ill-defined descriptions of heart disease); I52 (other heart disorders in diseases classified elsewhere). |
|  | **Pericardial diseases** | Diagnosis  (ICD-9) 391 (chronic rheumatic pericarditis); 420.xx (acute pericarditis); 423.x (other diseases of the pericardium);  (ICD-10) I30.x (acute pericarditis); I31.x (other diseases of the pericardium); I32 (pericarditis in diseases classified elsewhere) |
|  | **Valvular heart diseases** | Diagnosis  (ICD-9) 391.x (rheumatic fever with heart involvement); 394.x (diseases of the mitral valve); 395.x (diseases of the aortic valve); 396.x (diseases of the mitral and aortic valve); 397.x (diseases of other endocardial structures); 421.x (acute and subacute endocarditis); 424.xx (other diseases of the endocardium);  (ICD-10) I01.x (acute rheumatic fever w/ heart involvement); I05.x (rheumatic mitral valve disease); I06.x (rheumatic aortic valve disease); I07.x (rheumatic tricuspid valve disease); I08.x (multiple valve disease); I09.x (other rheumatic heart diseases); I33.x (acute and subacute endocarditis); I34.x (nonrheumatic mitral valve disorders); I35.x (nonrheumatic aortic valve disorders); I36.x (nonrheumatic tricuspid valve disorders); I37.x (nonrheumatic pulmonary valve disorders); I38 (endocarditis, valve unspecified); I39 (endocarditis and heart valve disorders in diseases classified elsewhere) |
|  | **Arrhythmias** | Diagnosis  (ICD-9) 426.xx (conduction disorders); 427.xx (cardiac dysrhythmias)  (ICD-10) I44.xx (atrioventricular and left bundle branch block); I45.xx (other conduction disorders); I47.x (paroxysmal tachycardia); I48.xx (atrial fibrillation and flutter); I49.xx (other cardiac arrhythmias.  Procedure  (ICD-10) 02K8xxx (conduction mapping) |
|  | **Aortic Disease** | Diagnosis  (ICD-9) 441.xx (aortic aneurysm and dissection), (ICD-10) I71.xx (aortic aneurysm and dissection) |
|  | **Congenital heart disease** | Diagnosis  (ICD-9) 745.xx (bulbus cordis anomalies and anomalies of cardiac septal closure); 746.xx (other congenital anomalies of heart); 747.0x - 747.4x (other congenital anomalies of the circulatory system).  (ICD-10) Q20.x (congenital malformations of cardiac chambers and connections); Q21.x (congenital malformations of the cardiac septa); Q22.x (congenital malformations of the pulmonary and tricuspid valves); Q23.x (congenital malformations of the aortic and mitral valves); Q24.x (other congenital malformations of the heart); Q25.xx (congenital malformations of great arteries); Q26.x (congenital abnormalities of great veins) |
|  | **Other CAD** | Diagnosis  (ICD-10) I25.4x (coronary artery aneurysm and dissection) |
| No CVD | No codes for aCVD or naCVD as defined above (if 1 code for aCVD or naCVD they would be excluded from the study as it is uncertain which group they fall into). | |
| Diabetes | Diagnosis  (ICD-9) 249.xx (secondary diabetes); 250.xx (diabetes mellitus); 357.2 (diabetic polyneuropathy); 362.0x (diabetic retinopathy); 366.41 (diabetic cataract)  (ICD-10) E08.x (diabetes mellitus due to underlying condition), E09.x (drug or chemical-induced diabetes mellitus), E10.x (type 1 diabetes mellitus), E11.x (type 2 diabetes mellitus), E13.x (other specified diabetes mellitus) E14.x (Unspecified diabetes mellitus) | |
| Hypertension | Diagnosis  (ICD-9) 401.x (essential hypertension); 403.xx (hypertensive chronic kidney disease); 405.xx (secondary hypertension)  (ICD-10) I10 (primary hypertension); I12.x (hypertensive chronic kidney disease); I15.x (secondary hypertension) | |
| Obesity | BMI as provided in MarketScan data (continuous variable) | |
| Chronic Kidney Disease | Diagnosis  (ICD-9) 585.3, 585.4  (ICD-10) N18.3x (CKD stage 3); N18.4 (CKD stage 4); N18.5 (CKD stage 5); N18.6 (ESRD) | |
| Hyperlipidemia | Diagnosis  (ICD-9) 272  (ICD-10) E78.xx (disorders of lipoprotein metabolism, excluding E78.7 and E78.8); E88.81 (metabolic syndrome) | |
| Tobacco use | MarketScan data, self-reported: Currently using cigars (yes/no), previously used cigars (yes/no), number of cigarettes smoked per day (coded 1-4 based on response), currently using cigarettes (yes/no), number of years of cigarette use (coded 1-4 based on response), packs of cigarettes daily (<1, >1), previous use of cigarettes (yes/no), previous use of chewing tobacco (yes/no), current use of chewing tobacco (yes/no) | |
| **Cancer diagnosis** |  | |
| Lip, oral cavity, pharynx (head and neck) | Diagnosis  (ICD-9) 140.0, 140.1, 140.3, 140.4, 140.5, 140.6, 140.8, 140.9, 141.0, 141.1, 141.2, 141.3, 141.4, 141.5, 141.6, 141.8, 141.9, 142.0, 142.1, 142.2, 142.8, 142.9, 143.0, 143.1, 143.8, 143.9, 144.0, 144.1, 144.8, 144.9, 145.0, 145.1, 145.2, 145.3, 145.4, 145.5, 145.6, 145.8, 145.9, 146.0, 146.1, 146.2, 146.3, 146.4, 146.5, 146.6, 146.7, 146.8, 146.9, 147.0, 147.1, 147.2, 147.3, 147.8, 147.9, 148.0, 148.1, 148.2, 148.3, 148.8, 148.9, 149.0, 149.1, 149.8, 149.9  (ICD-10) C00.0, C00.1, C00.2, C00.3, C00.4, C00.5, C00.6, C00.7, C00.8, C00.9, C01, C02.0, C02.1, C02.2, C02.3, C02.4, C02.8, C02.9, C03.0, C03.1, C03.9, C04.0, C04.1, C04.8, C04.9, C05.0, C05.1, C05.2, C05.8, C05.9, C06.0, C06.1, C06.2, C06.8, C06.9, C07, C08.0, C08.1, C08.9, C09.0, C09.1, C09.8, C09.9, C10.0, C10.1, C10.2, C10.3, C10.4, C10.8, C10.9, C11.0, C11.1, C11.2, C11.3, C11.8, C11.9, C12, C13.0, C13.1, C13.2, C13.8, C13.9, C14.0, C14.2, C14.8 | |
| Esophagus | Diagnosis (ICD-9) 150.0, 150.1, 150.2, 150.3, 150.4, 150.5, 150.8, 150.9  (ICD-10) C15.3, C15.4, C15.5, C15.8, C15.9 | |
| Stomach | Diagnosis (ICD-9) 151.0, 151.1, 151.2, 151.3, 151.4, 151.5, 151.6, 151.8, 151.9  (ICD-10) C16.0, C16.1, C16.2, C16.3, C16.4, C16.5, C16.6, C16.8, C16.9 | |
| Small intestine | Diagnosis (ICD-9) 152.0, 152.1, 152.2, 152.3, 152.8, 152.9  (ICD-10) C17.0, C17.1, C17.2, C17.3, C17.8, C17.9, C26.0 | |
| Colon | Diagnosis (ICD-9) 153.0, 153.1, 153.2, 153.3, 153.4, 153.5, 153.6, 153.7, 153.8, 153.9  (ICD-10) C18.0, C18.1, C18.2, C18.3, C18.4, C18.5, C18.6, C18.7, C18.8, C18.9 | |
| Rectosigmoid, rectum, anus | Diagnosis (ICD-9) 154.0, 154.1, 154.2, 154.3, 154.8  (ICD-10) C19, C20, C21.0, C21.1, C21.2, C21.8 | |
| Liver, gallbladder, spleen | Diagnosis (ICD-9) 155.0, 155.1, 155.2, 156.0, 156.1, 156.2, 156.8, 156.9, 159.0, 159.1  (ICD-10) C22.0, C22.1, C22.2, C22.3, C22.4, C22.7, C22.8, C22.9, C23, C24.0, C24.1, C24.8, C24.9, C26.1, C26.9 | |
| Pancreas | Diagnosis (ICD-9) 157.0, 157.1, 157.2, 157.3, 157.4, 157.8, 157.9  (ICD-10) C25.0, C25.1, C25.2, C25.3, C25.4, C25.7, C25.8, C25.9 | |
| Nasal cavity, middle ear, accessory sinuses | Diagnosis (ICD-9) 160.0, 160.1, 160.2, 160.3, 160.4, 160.5, 160.8, 160.9  (ICD-10) C30.0, C30.1, C31.0, C31.1, C31.2, C31.3, C31.8, C31.9 | |
| Larynx, trachea | Diagnosis (ICD-9) 161.0, 161.1, 161.2, 161.3, 161.8, 161.9, 162.0  (ICD-10) C32.0, C32.1, C32.2, C32.3, C32.8, C32.9, C33 | |
| Lung | Diagnosis (ICD-9) 162.2, 162.3, 162.4, 162.5, 162.8, 162.9, 165.0, 165.8, 165.9  (ICD-10) C34.0, C34.1, C34.2, C34.3, C34.8, C34.9, C39.0, C39.9 | |
| Thymus, heart, mediastinum, pleura | Diagnosis (ICD-9) 163.0, 163.1, 163.8, 163.9, 164.0, 164.1, 164.2, 164.3, 164.8, 164.9  (ICD-10) C37, C38.0, C38.1, C38.2, C38.3, C38.4, C38.8 | |
| Bone and articular cartilage | Diagnosis (ICD-9) 170.0, 170.1, 170.2, 170.3, 170.4, 170.5, 170.6, 170.7, 170.8, 170.9  (ICD-10) C40.0, C40.1, C40.2, C40.3, C40.8, C40.9, C41.0, C41.1, C41.2, C41.3, C41.4, C41.9 | |
| Melanoma | Diagnosis (ICD-9) 172.0, 172.1, 172.2, 172.3, 172.4, 172.5, 172.6, 172.7, 172.8, 172.9  (ICD-10) C43.0, C43.1, C43.2, C43.3, C43.4, C43.5, C43.6, C43.7, C43.8, C43.9 | |
| Mesothelial and soft tissue | Diagnosis (ICD-9) 158.0, 158.8, 158.9, 171.0, 171.2, 171.3, 171.4, 171.5, 171.6, 171.7, 171.8, 171.9, 176.0, 176.1, 176.2, 176.3, 176.4, 176.5, 176.8, 176.9  (ICD-10) C45.0, C45.1, C45.2, C45.7, C45.9, C46.0, C46.1, C46.2, C46.3, C46.4, C46.5, C46.7, C46.9, C47.0, C47.1, C47.2, C47.3, C47.4, C47.5, C47.6, C47.8, C47.9, C48.0, C48.1, C48.2, C48.8, C49.0, C49.1, C49.2, C49.3, C49.4, C49.5, C49.6, C49.8, C49.9, C49.A | |
| Breast | Diagnosis (ICD-9) 174.0, 174.2, 174.3, 174.4, 174.5, 174.6, 174.8, 174.9, 175.0, 175.9  (ICD-10) C50.0, C50.1, C50.2, C50.3, C50.4, C50.5, C50.6, C50.8, C50.9 | |
| Uterine | Diagnosis (ICD-9) 179, 182.0, 182.1, 182.8  (ICD-10) C54.0, C54.1, C54.2, C54.3, C54.8, C54.9, C55 | |
| Ovarian | Diagnosis (ICD-9) 183  (ICD-10) C56.1, C56.2, C56.9 | |
| Female genital organs, other | Diagnosis (ICD-9) 180.0, 180.1, 180.8, 180.9, 181, 183.2, 183.3, 183.4, 183.5, 183.8, 183.9, 184.0, 184.1, 184.2, 184.3, 184.4, 184.8, 184.9  (ICD-10) C51.0, C51.1, C51.2, C51.8, C51.9, C52, C53.0, C53.1, C53.8, C53.9, C57.0, C57.1, C57.2, C57.3, C57.4, C57.7, C57.8, C57.9, C58 | |
| Prostate | Diagnosis (ICD-9) 185  (ICD-10) C61, Z19.1, Z19.2 | |
| Male genital organs, other | Diagnosis (ICD-9) 186.0, 186.9, 187.1, 187.2, 187.3, 187.4, 187.5, 187.6, 187.7, 187.8, 187.9  (ICD-10) C60.0, C60.1, C60.2, C60.8, C60.9, C62.0, C62.1, C62.9, C63.0, C63.1, C63.2, C63.7, C63.8, C63.9 | |
| Renal cell carcinoma | Diagnosis (ICD-9) 189.0, 189.1  (ICD-10) C64.1, C64.2, C64.9, C65.1, C65.2, C65.9, C66.1, C66.2, C66.9 | |
| Bladder | Diagnosis (ICD-9) 188.0, 188.1, 188.2, 188.3, 188.4, 188.5, 188.6, 188.7, 188.8, 188.9  (ICD-10) C67.0 , C67.1, C67.2, C67.3, C67.4, C67.5, C67.6, C67.7, C67.8, C67.9 | |
| Urinary organs, other | Diagnosis (ICD-9) 189.2, 189.3, 189.4, 189.8, 189.9  (ICD-10) C68.0, C68.1, C68.8, C68.9 | |
| Eye and adnexa | Diagnosis (ICD-9) 190.0, 190.1, 190.2, 190.3, 190.4, 190.5, 190.6, 190.7, 190.8, 190.9  (ICD-10) C69.0, C69.1, C69.2, C69.3, C69.4, C69.5, C69.6, C69.8, C69.9 | |
| Brain, meninges, nerves | Diagnosis (ICD-9) 191.0, 191.1, 191.2, 191.3, 191.4, 191.5, 191.6, 191.7, 191.8, 191.9, 192.0, 192.1, 192.2, 192.3, 192.8, 192.9  (ICD-10) C70.0, C70.1, C70.9, C71.0, C71.1, C71.2, C71.3, C71.4, C71.5, C71.6, C71.7, C71.8, C71.9, C72.0, C72.1, C72.2, C72.3, C72.4, C72.5, C72.9 | |
| Thyroid, endocrine (other) | Diagnosis (ICD-9) 193, 194.0, 194.1, 194.3, 194.4, 194.5, 194.6, 194.8, 194.9  (ICD-10) C73, C74.0, C74.1, C74.9, C75.0, C75.1, C75.2, C75.3, C75.4, C75.5, C75.8, C75.9 | |
| Neuroendocrine tumors | Diagnosis (ICD-9) 209.0, 209.1, 209.2, 209.3, 209.4, 209.5, 209.6, 209.7  (ICD-10) C7A.0, C7A.1, C7A.8, C7B.0, C7B.1, C7B.8 | |
| Lymphoma (B and T), plasma cell dyscrasias | Diagnosis (ICD-9) 200.0, 200.1, 200.2, 200.3, 200.4, 200.5, 200.6, 200.7, 200.8, 201.0, 201.1, 201.2, 201.4, 201.5, 201.6, 201.7, 201.9, 202.0, 202.1, 202.2, 202.3, 202.4, 202.5, 202.7, 202.8, 202.9, 203.0, 203.1  (ICD-10) C81.0, C81.1, C81.2, C81.3, C81.4, C81.7, C81.9, C82.0, C82.1, C82.2, C82.3, C82.4, C82.5, C82.6, C82.8, C82.9, C83.0, C83.1, C83.3, C83.5, C83.7, C83.8, C83.9, C84.0, C84.1, C84.4, C84.6, C84.7, C84.A, C84.Z, C84.9, C85.1, C85.2, C85.8, C85.9, C86.0, C86.1, C86.2, C86.3, C86.4, C86.5, C86.6, C88.0, C88.2, C88.3, C88.4, C88.8, C88.9, C90.0, C90.1, C90.2, C90.3 | |
| Leukemia (lymphoid, myeloid, and other) | Diagnosis (ICD-9) 204.0, 204.1, 204.2, 204.8, 204.9, 205.0, 205.1, 205.2, 205.3, 205.8, 205.9, 206.0, 206.1, 206.2, 206.8, 206.9, 207.0, 207.1, 207.2, 207.8, 208.0, 208.1, 208.2, 208.8, 208.9  (ICD-10) C91.0, C91.1, C91.3, C91.4, C91.5, C91.6, C91.A, C91.Z, C91.9, C92.0, C92.1, C92.2, C92.3, C92.4, C92.5, C92.6, C92.A, C92.Z, C92.9, C93.0, C93.1, C93.3, C93.Z, C93.9, C94.0, C94.2, C94.3, C94.4, C94.6, C94.8, C95.0, C95.1, C95.9 | |
| Hematologic malignancies, other | Diagnosis (ICD-9) 202.6, 203.8, 238.4, 238.7  (ICD-10) C96.0, C96.2, C96.4, C96.5, C96.6, C96.A, C96.Z, C96.9, D45, D46 | |
| Personal history of malignancy (only used for exclusion but not for incident cancer) | Diagnosis (ICD-9) V10.x (excluding V10.83), V87.41, V87.43  (ICD-10) Z85.x (excluding Z85.82), Z86.00x, Z92.21, Z92.23 | |

ICD, International Classification of Disease; CM, Clinical Modification; PCS, Procedure Coding System; CPT, Current Procedural Terminology; CVD, cardiovascular disease; STEMI, ST-elevation myocardial infarction; NSTEMI, non-ST-elevation myocardial infarction; PCI, percutaneous coronary intervention; PCTA, percutaneous transluminal coronary angioplasty; CABG, coronary artery bypass grafting; CKD, chronic kidney disease; ESRD, end-stage renal disease.

**Table 2. Baseline characteristics of 1:1 matched cohorts by cardiovascular disease (CVD) group**

|  | **No CVD** | **aCVD** | **naCVD** |
| --- | --- | --- | --- |
|  | **(N=2,243,706)** | **(N=1,388,981)** | **(N=854,725)** |
|  | **N (%)** | **N (%)** | **N (%)** |
| Age at first enrollment |  |  |  |
| Mean(std) | 54 (17) | 62 (13) | 52 (16) |
| Median (IQR) | 56 (45, 65) | 60 (53, 71) | 53 (42, 61) |
| 18-39 | 428348 (19.1) | 53944 (3.9) | 183515 (21.5) |
| 40-49 | 327274 (14.6) | 158769 (11.4) | 176286 (20.6) |
| 45-59 | 616422 (27.5) | 436311 (31.4) | 232347 (27.2) |
| 60-64 | 279451 (12.5) | 210651 (15.2) | 88055 (10.3) |
| 65-69 | 182814 (8.1) | 151606 (10.9) | 53371 (6.2) |
| 70-79 | 281483 (12.5) | 248080 (17.9) | 81830 (9.6) |
| ≥80 | 127914 (5.7) | 129620 (9.3) | 39321 (4.6) |
| Female | 1075928 (48) | 600668 (43.2) | 475260 (55.6) |
| Diabetes | 549814 (24.5) | 415209 (29.9) | 155696 (18.2) |
| Hypertension | 1471549 (65.6) | 956875 (68.9) | 514674 (60.2) |
| CKD | 63862 (2.8) | 43253 (3.1) | 16851 (2) |
| Hyperlipidemia | 1407731 (62.7) | 927649 (66.8) | 480082 (56.2) |
| Region |  |  |  |
| Northeast | 462972 (20.6) | 347265 (25) | 221148 (25.9) |
| North central | 530317 (23.6) | 349695 (25.2) | 179883 (21) |
| South | 802216 (35.8) | 486786 (35) | 306657 (35.9) |
| West | 393225 (17.5) | 170515 (12.3) | 126760 (14.8) |
| Unknown | 54976 (2.5) | 34720 (2.5) | 20277 (2.4) |
| Insurance |  |  |  |
| PPO | 1244271 (55.5) | 749678 (54) | 496818 (58.1) |
| HMO | 321499 (14.3) | 164724 (11.9) | 105337 (12.3) |
| Other | 552088 (24.6) | 390342 (28.1) | 199419 (23.3) |
| Unknown | 125848 (5.6) | 84237 (6.1) | 53151 (6.2) |

Abbreviations: CVD, cardiovascular disease; naCVD, non-atherosclerotic cardiovascular disease; aCVD, atherosclerotic cardiovascular disease; IQR, interquartile range; CKD, chronic kidney disease; PPO, preferred provider organization; HMO, health maintenance organization.

**Table 3.** **Baseline characteristics of HRA-linked cohorts by cardiovascular (CVD) group** (N=1,282,261)

|  | **No CVD** | **CVD** | **aCVD** | **naCVD** |
| --- | --- | --- | --- | --- |
|  | **(N=1,220,486)** | **(N=61,775)** | **(N=29,672)** | **(N=32,103)** |
|  | **N (%)** | **N (%)** | **N (%)** | **N (%)** |
| Age at first enrollment, |  |  |  |  |
| Median (range) | 40 (18,64) | 51 | 54 | 48 |
| 18-39 | 599958 (49.2) | 11120 (18) | 2500 (8.4) | 8620 (26.9) |
| 40-49 | 330873 (27.1) | 15640 (25.3) | 6802 (22.9) | 8838 (27.5) |
| 45-59 | 255920 (21) | 27773 (45) | 15710 (52.9) | 12063 (37.6) |
| 60-64 | 33735 (2.8) | 7242 (11.7) | 4660 (15.7) | 2582 (8) |
| Female | 635352 (52.1) | 24970 (40.4) | 9722 (32.8) | 15248 (47.5) |
| Self-reported smoker | 212880 (17.4) | 12308 (19.9) | 6863 (23.1) | 5445 (17) |
| Self-reported Body Mass Index |  |  |  |  |
| Light / normal (<25 kg/m^2^)) | 334006 (27.4) | 10281 (16.6) | 3685 (12.4) | 6596 (20.5) |
| Overweight ((≥25 but <30 kg/m^2^) | 360091 (29.5) | 16654 (27) | 7911 (26.7) | 8743 (27.2) |
| Obese (≥30kg/m^2^) | 321343 (26.3) | 20703 (33.5) | 10279 (34.6) | 10424 (32.5) |
| Unknown | 205046 (16.8) | 14137 (22.9) | 7797 (26.3) | 6340 (19.7) |
| Diabetes | 77489 (6.3) | 11599 (18.8) | 7536 (25.4) | 4063 (12.7) |
| Hypertension | 224461 (18.4) | 31354 (50.8) | 17958 (60.5) | 13396 (41.7) |
| CKD | 2895 (0.2) | 1028 (1.7) | 636 (2.1) | 392 (1.2) |
| Hyperlipidemia | 280359 (23) | 32961 (53.4) | 19742 (66.5) | 13219 (41.2) |
| Region |  |  |  |  |
| Northeast | 153267 (12.6) | 7839 (12.7) | 3359 (11.3) | 4480 (14) |
| North central | 369028 (30.2) | 20462 (33.1) | 10545 (35.5) | 9917 (30.9) |
| South | 466037 (38.2) | 24492 (39.6) | 11849 (39.9) | 12643 (39.4) |
| West | 208551 (17.1) | 7728 (12.5) | 3304 (11.1) | 4424 (13.8) |
| Unknown | 23603 (1.9) | 1254 (2) | 615 (2.1) | 639 (2) |
| Insurance |  |  |  |  |
| PPO | 676451 (55.4) | 35702 (57.8) | 17273 (58.2) | 18429 (57.4) |
| HMO | 179510 (14.7) | 9450 (15.3) | 4309 (14.5) | 5141 (16) |
| Other | 337231 (27.6) | 15185 (24.6) | 7377 (24.9) | 7808 (24.3) |
| Unknown | 27294 (2.2) | 1438 (2.3) | 713 (2.4) | 725 (2.3) |

Abbreviations: HRA, health risk assessment; kg, kilogram; CVD, cardiovascular disease; aCVD, atherosclerotic cardiovascular disease; naCVD, non-atherosclerotic cardiovascular disease; CKD, chronic kidney disease; PPO, preferred provider organization; HMO, health maintenance organization.

Note: All P-values were significant <0.001.

**Table 4. Cox proportional hazards model of time to cancer for HRA-linked data** (N=1,282,261)

|  | **Hazard Ratio** | **95% CI** | **P** |
| --- | --- | --- | --- |
| *No weight adjustment, no propensity score adjustment* | | | |
| No CVD vs. |  |  |  |
| CVD | 1.16 | 1.11 - 1.21 | <0.001 |
| *No weight adjustment, no propensity score adjustment* | | | |
| No CVD vs. |  |  |  |
| naCVD | 1.13 | 1.06 - 1.20 | <0.001 |
| aCVD | 1.19 | 1.12 - 1.25 | <0.001 |
| naCVD vs. |  |  |  |
| No CVD | 0.89 | 0.84 - 0.94 | <0.001 |
| aCVD | 1.05 | 0.98 - 1.14 | 0.18 |
| *Propensity score adjustment, no weight adjustment* | | | |
| No CVD vs. |  |  |  |
| CVD | 1.16 | 1.11 - 1.21 | <0.001 |
| *Propensity score adjustment, no weight adjustment* | | | |
| No CVD vs. |  |  |  |
| naCVD | 1.13 | 1.06 - 1.20 | <0.001 |
| aCVD | 1.18 | 1.12 - 1.25 | <0.001 |
| naCVD vs. |  |  |  |
| No CVD | 0.89 | 0.84 - 0.94 | <0.001 |
| aCVD | 1.05 | 0.97 - 1.14 | 0.20 |
| *Weight adjustment, weights truncated at 5% or 95%* | | | |
| No CVD vs. |  |  |  |
| naCVD | 1.12 | 0.93 - 1.35 | 0.25 |
| aCVD | 1.15 | 0.98 - 1.34 | 0.08 |
| naCVD vs. |  |  |  |
| No CVD | 0.9 | 0.74 - 1.08 | 0.25 |
| aCVD | 1.03 | 0.82 - 1.29 | 0.82 |

Abbreviations: HRA, health risk assessment; CI, confidence interval. CVD, cardiovascular disease; naCVD, non-atherosclerotic cardiovascular disease; aCVD, atherosclerotic cardiovascular disease.

Models were additionally adjusted for first enrollment year, age, sex, baseline diabetes, chronic kidney disease, hyperlipidemia, self-reported smoking status and Body Mass Index, region, and insurance.
